# The interaction between lifetime depression severity and BMI is related to altered activation pattern in the right inferior frontal gyrus during food anticipation

**DOI:** 10.1101/2021.02.17.21251922

**Authors:** A Manelis, YO Halchenko, S Satz, R Ragozzino, M Lucero, HA Swartz, MD Levine

**Affiliations:** Department of Psychiatry, University of Pittsburgh, Pittsburgh, PA; Department of Psychological and Brain Sciences, Dartmouth College, NH, USA

**Keywords:** depression, BMI, overweight/obese, fMRI, anticipation, food, right inferior frontal gyrus

## Abstract

**Background:** Depression and obesity often co-occur but the underlying neural mechanisms for this bidirectional link are not well understood. Using fMRI, we examined how the relationship between Body Mass Index (BMI) and dimensional lifetime depression severity was associated with brain activation during food anticipation and pleasant/unpleasant rating.

**Methods:** 90 participants (48 healthy controls, 42 with unipolar depression (UD), 69 female, age=28.5±6.6) were fMRI-scanned while performing the Food and Object Cued Encoding task consisting of food/object anticipation and food/object pleasant/unpleasant rating phases.

**Results:** The analysis across all participants revealed a significant BMI-by-lifetime depression severity interaction on RIFG activation during food anticipation (p<0.0125). Most symptomatic and overweight/obese individuals with UD showed decreased right inferior frontal gyrus (RIFG) activation during food anticipation, while less symptomatic and/or normal-weight individuals with UD showed increased RIFG activation during food anticipation. RIFG activation during food anticipation was negatively correlated with RIFG activation during pleasant/unpleasant rating (r= -0.63, p<0.001). Individuals with UD who showed higher RIFG activation for food items during pleasant/unpleasant rating reported liking or wanting those food items less than those with lower RIFG activation (p<0.05).

**Conclusions:** The IFG is involved in emotion regulation and response inhibition necessary to control appetitive behavior. Greater RIFG activation during pleasant/unpleasant rating of food coupled with low ratings of food liking and wanting could be associated with inhibition of cognitive and emotional response to food in UD. This process may be cognitively challenging and stressful thus putting affected individuals with UD at risk for weight gain and worsening of depression.

## 1 INTRODUCTION

Based on the World Health Organization fact sheet, over 1.9 billion adults are overweight, of which over 650 million are overweight/obese (https://www.who.int/news-room/fact-sheets/detail/obesity-and-overweight). Taken together with the fact that over 264 million people worldwide suffer from depression (https://www.who.int/news-room/fact-sheets/detail/depression), it becomes clear that both unipolar depression (UD) (1) and obesity (2) are important personal and public health problems. UD and overweight/obesity often co-occur (3) and may share common biological pathways explaining the bidirectional link between them (4). For example, being overweight/obese increased the risk of developing mood and anxiety disorders (5,6), while losing weight was associated with the depression severity improvements (7,8). The link between depression and obesity may be explained by biological and psychological factors including genetics, functioning of systems involved in homeostatic adjustments (9), and brain networks integrating homeostatic responses and emotion regulation (4).

Recent large-scale studies of brain structure found lower grey matter volume in obese vs. normal-weight individuals (10,11). Functional magnetic resonance studies (fMRI) showed that obese, compared to normal weight, individuals have greater activation in the left dorsomedial prefrontal, right inferior frontal, superior frontal, anterior cingulate cortices, and parahippocampal gyri, but lower activation in the left dorsolateral prefrontal and insular cortices for food vs non-food stimuli (for meta-analysis see (12)). Neuroimaging studies comparing individuals with depression vs. healthy controls (HC) identified aberrant activation in the anterior cingulate cortex, insula, amygdala, prefrontal cortex, and striatum among other regions in response to negative emotional stimuli (13,14).

Although understanding the neurobiological factors contributing to the interplay between depression and obesity is critical for development of prevention and intervention treatments, to date, only one neuroimaging study has focused on comparing individuals with obesity vs. those with obesity and major depressive disorder (15). That study found that individuals with obesity and depression had stronger activation in the precuneus and anterior cingulate cortex compared to obese individuals without depression during pleasant/unpleasant judgments of words. These findings suggest that both obesity and depression are associated with aberrant functioning of brain systems supporting reward processing, executive function (e.g., response inhibition) and cognitive control.

It was proposed that in healthy individuals, hunger may increase motivational and neural processing of food-related cues during the expectation of food-related stimuli thus influencing hedonic-driven food consumption (16). For example, ventral striatal activation during anticipation of food reward positively correlated with Body Mass Index (BMI) (17) and was higher for food-related reward for hungry vs. satiated participants (16). No study so far examined neural underpinnings of food *anticipation* when no food cues were available in overweight/obese vs. normal weight individuals with and without lifetime history of depression. Considering that previous research in our lab found that HC could be distinguished from individuals with mood disorders based on brain activation during *anticipation* of emotional stimuli (18,19), we propose that the depression-obesity interaction may affect processing of food-related anticipatory cues. To test this hypothesis, we examined neural correlates underlying the interaction between dimensional life-time depression severity and BMI during a task that involved anticipation of food items and then, judging food pictures as pleasant or unpleasant. We aimed to develop an empirical model that explains how the interplay between depression and obesity affects anticipatory and task-related brain activation for food stimuli and how this brain response is related to how much participants like or want to eat food items presented in the study.

## 2 METHOD

### 2.1 Participants

The study was approved by the University of Pittsburgh Institutional Review Board. Participants were recruited from the community, local universities, and medical centers. Written informed consent was obtained from all participants. Participants were right-handed, fluent in English, and matched on age, sex, and BMI. HC had no personal or family history of psychiatric disorders. Symptomatic individuals met DSM-5 criteria for depressive disorders such as major depressive or persistent depressive disorders referred here as ‘unipolar depression’ (UD). We scanned 101 participants (53 HC and 48 UD). Of them, 6 HC and 5 UD were removed from the analyses due to excessive motion (>4mm between the fMRI volumes in any direction), scanning artifacts, or more than 20% of missing responses on the task, thus leaving 90 participants (47 HC and 43 UD) in the analyses.

### 2.2. Clinical assessment

All diagnoses were made by a trained clinician and confirmed by a psychiatrist according to DSM-5 criteria using SCID-5 (20). We also assessed the current depression symptoms (Hamilton Depression Rating Scale (HDRS-25) (21)), current mania symptoms (Young Mania Rating Scale YMRS (22)), and lifetime dimensional symptoms of depression (Moods Spectrum self-report questionnaire MOODS-SR (23)). A total psychotropic medication load was calculated for each participant. Greater numbers and doses of medications corresponding to a greater medication load (19,24). Exclusion criteria included a history of head injury, metal in the body, pregnancy, claustrophobia, neurodevelopmental disorders, systemic medical illness, premorbid IQ<85 per the National Adult Reading Test (NART; (25)), current alcohol/drug abuse, YMRS scores>10 at scan, meeting criteria for any psychotic-spectrum disorder. Table 1 reports group statistics for participants’ demographic and clinical characteristics.

**Table 1.** Demographic and clinical characteristics

|  | HC | UD | statistics HC vs. UD |
| --- | --- | --- | --- |
| N | 48 | 42 |  |
| Gender (number females) | 36 | 33 | chi2=0.02, p=0.88 |
| UD diagnoses (MDD/PDD) | na | 26/16 | na |
| Age (years) | 28.09(0.91) | 28.98(1.08) | t(88)=-0.64,p=0.52 |
| BMI | 25.82(0.65) | 25.73(0.67) | t(88)=0.1,p=0.92 |
| Number of overweight/obese participants (total %) | 15/8 (48%) | 13/8 (50%) | ns |
| IQ (NART) | 106.72(0.82) | 110.64(1.08) | t(88)=-2.92,p<0.01 |
| VAS before scan | 24.18(2.76) | 22.85(3.5) | t(55)=0.3,p=0.76 |
| Current depression severity (HRSD-25) | 1.75(0.3) | 13.52(1.1) | t(88)=-10.92,p<0.001 |
| Lifetime depression (MOODS-SR) | 2.1(0.33) | 18.71(0.66) | t(88)=-23.22,p<0.001 |
| Illness Onset (year of age) | na | 15.05(0.76) | na |
| Number of participants taking Antidepressants | na | 27 | na |
| Number of participants taking Mood stabilizers | na | 3 | na |
| Number of participants taking Antipsychotics | na | 1 | na |
| Number of participants taking Benzodiazepines | na | 3 | na |
| Number of participants taking Stimulants | na | 4 | na |
| A mean number of psychotropic medications | na | 1.14(0.16) | na |
| A mean total medication load | na | 1.48(0.22) | na |
| Number of participants with comorbid diagnoses | na | 29 | na |

Note. The table reports the mean and standard error of mean (SE) in parenthesis. ns - not significant; na - not applicable.

### 2.3 Behavioral assessments

Participants rated their feelings towards food (i.e., hunger, fullness, urge to eat) before the scan using a 6-question Visual Analogues Scale (VAS) ranging from 0 (‘Not at all’) to 100 (‘Extremely’). These data were only available for 33 HC and 24 UD.

Inside the scanner, participants performed a Food and Object Cued Encoding Task (Fig.1). Each trial of this task started with the 4-second *anticipation* phase during which participants were presented with either a triangle predicting food or a circle predicting object categories of pictures. Participants were instructed to mentally prepare to process the category of items predicted by the cue. After the cue, a stimulus from the predicted category taken from the *Food-pics* image database (26) was shown (maximum duration=1.5sec) and participants rated the stimulus as pleasant or unpleasant (the *pleasant/unpleasant rating*) by pressing a corresponding button with the index finger on one hand for pleasant images and on the other hand for unpleasant images. The hand assignment for pleasant/unpleasant responses was counterbalanced across subjects. A total of 48 trials (10-11 seconds each) were presented over two 4-minute runs.

After the scan, the participants were shown the food pictures that they saw during the scan and were asked to indicate on a 9-point scale (1 – do not like/want it, 9 – like/want it very much) how much they normally like to eat those items (‘LIKE’ condition) and how much they wanted to eat them ‘right now’ at the time of the assessment (‘WANT’ condition).

**Figure 1.**
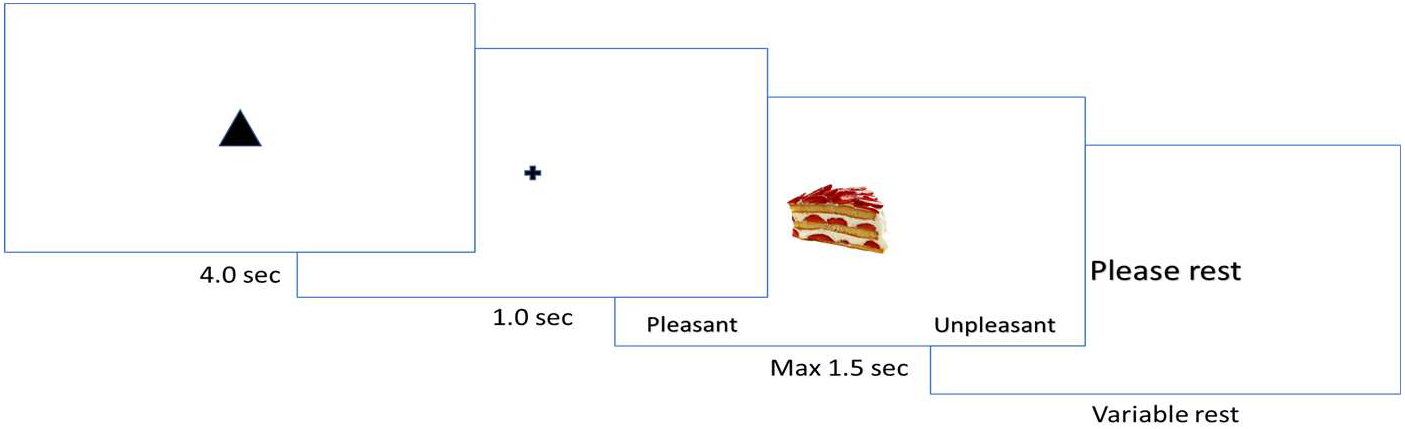
An example of a trial in the Food and Object Cued Encoding Task.

### 2.4 Neuroimaging data acquisition

The neuroimaging data were collected at the University of Pittsburgh/UPMC Magnetic Resonance Research Center using a 3T Siemens Prisma scanner with a 64-channel coil and named according to the ReproIn convention (27). The EPI data were collected in the anterior-to-posterior direction using a multi-band sequence (factor=8, TR=800ms, resolution=2×2×2mm, FOV=210, TE=30ms, flip angle=52°, 72 slices, 315 volumes). High-resolution T1w images were collected using the MPRAGE sequence (TR=2400ms, resolution=0.8×0.8×0.8mm, 208 slices, FOV=256, TE=2.22ms, flip angle=8°). Field maps were collected in the AP and PA directions using the spin echo sequence (TR=8000, resolution=2×2×2mm, FOV=210, TE=66ms, flip angle=90°, 72 slices).

### 2.5 Data analyses

#### 2.5.1 Clinical data analysis

The demographic and clinical characteristics were compared between groups using t- and chi-square tests. Partial correlation analysis (R package *ppcor* (28)) examined the relationship between lifetime depression severity and BMI as well as the relationship between lifetime depression severity and ‘LIKE’ and ‘WANT’ ratings while accounting for age, sex and IQ separately in HC and UD.

#### 2.5.2 Behavioral data analysis

The available VAS data were compared between groups using a t-test. A percent of ‘pleasant’ responses and RT were analyzed using a mixed effects model (‘lme4’(29), ‘lmerTest’(30), and ‘psycho’(31) packages in R) with group (UD/HC) as a between-subject factor, and condition (food/objects) as a within-subject factor.

#### 2.5.3 Neuroimaging data analysis

##### Preprocessing

The DICOM images were converted to NIFTI and bids dataset using *heudiconv* (32). Data quality was examined using *mriqc* 0.15.1 (33). The data were preprocessed using *fmriprep* 20.1.1 (34). Preprocessing steps are described in detail in Supplemental Materials and include the boilerplate text automatically generated by *fmriprep*. In short, T1w images were skull-stripped, brain surfaces were reconstructed using recon-all (FreeSurfer 6.0.1) (35), and brain masks were generated. For each BOLD, we applied motion correction, spatiotemporal filtering using *mcflirt* (36), and slice-timing correction using *3dTshift* (37). Two spin-echo images with opposing phase-encoding directions were used to correct for geometric distortion and improve co-registration. Preprocessing also included automatic removal of motional artifacts using ICA-AROMA (38) and spatial smoothing with an isotropic, Gaussian kernel of 6mm FWHM (full-width half-maximum), and regressing out non-steady state volumes. High-pass temporal filter (90-sec cutoff) was applied on the *fmriprep* preprocessed files.

##### Subject-level analysis

Subject-level statistical maps were computed using FSL 6.0.3 installed system-wide on the workstation with GNU/Linux Debian 10 operating system with NeuroDebian repository (39). A hemodynamic response was modeled using a gamma function. A subject-level model included 4 explanatory variables: food cues, object cues, food pictures, and object pictures. To account for individual differences in anticipatory processing as well as visual stimuli perception and processing, brain activation during *anticipation* and *pleasant/unpleasant rating* of objects was used as the baseline for the analyses of *anticipation* and *pleasant/unpleasant rating* of food pictures. The contrasts of interest included comparing food vs. object *anticipation* and food vs. object *pleasant/unpleasant rating*. Positive differences (increases) show greater activation for food vs. objects (food>objects), while negative differences (decreases) show lower activation for food vs. objects (food<objects).

##### Group-level analysis

First, we identified the brain regions that increased in activation either during food/object anticipation, or food/object picture processing, or both using the Sandwich Estimator (*swe*) approach (40) for nonparametric permutation inference that was run across all subjects using Threshold-Free Cluster Enhancement correction (TFCE) (41), 5000 permutations, and FWE-corrected p-values< 0.05 in the whole-brain mask. The resulting images were added to create an inclusive mask of voxels whose activation increased either during *anticipation* of food or objects, or during *pleasant/unpleasant rating* of food or object items, or during both.

Second, we identified brain regions within the inclusive mask described above whose activation was sensitive to BMI-by-lifetime depression severity interaction during anticipation and processing of food vs. object stimuli. Modelling dimensional lifetime depression instead of the UD/HD diagnostic status was chosen to capture both between-group and within-group variability. We again used the *swe* approach described above. The FWE-corrected p-values threshold for this analysis was setup to p=0.0125 (or 0.05/4) to Bonferroni correct for the two conditions of interest (*anticipation* and *pleasant/unpleasant rating*) and two contrasts (activation increase/decrease). Age, gender, and IQ were used as covariates. Functional localization was determined using the Harvard-Oxford cortical and subcortical structural atlases. The volumetric results were visualized on the brain surface using BrainNetViewer software (42).

##### Exploratory analyses

A partial correlation analysis (*ppcor* (28)) examined the relationship between activation for food vs. objects during *anticipation* and during *pleasant/unpleasant rating* in the regions identified in the group analysis described in the step two while accounting for age, sex, and IQ separately in HC and UD groups.

We also explored whether the UD/HC diagnostic status moderated the relationship between food-object *anticipation* or food-object *pleasant/unpleasant rating* and participant’s mean ‘LIKE’ and ‘WANT’ responses collected post-scan. The significance level for these analyses was Bonferroni corrected for the four moderator models: 0.05/4=0.0125.

We also explored the effect of medication load, age of illness onset, illness duration, number of episodes, and taking specific psychotropic medications on main findings in the UD group.

## 3 Results

### Demographic and Clinical

UD did not differ from HC in age, BMI, or gender composition, but had significantly higher IQ, and lifetime and current dimensional symptoms of depression (Table 1). When we controlled age, sex, and IQ on the relationship between BMI and lifetime depression severity, we found a significant positive partial correlation in UD (r=0.28, p=0.002), but not HC (r= -0.13, p=0.14). Higher BMI individuals with UD had more severe lifetime depression symptoms than their lower BMI counterparts (Figure 2). Hunger level and appetite did not differ between groups prior to scanning (p=0.76)

**Figure 2.**
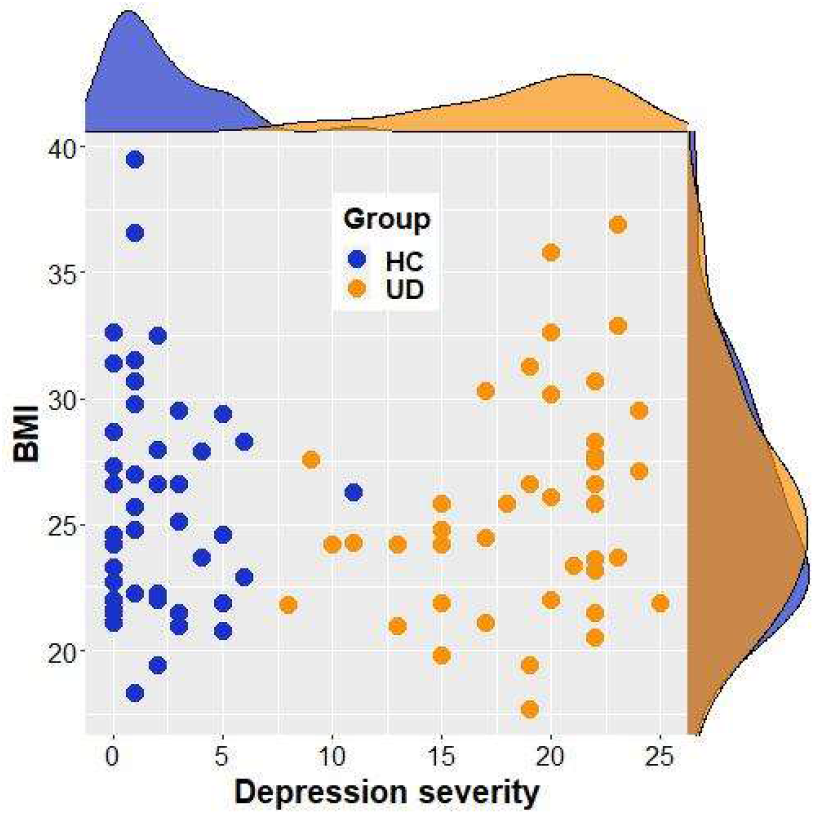
Lifetime depression severity (per MOODs-SR depression scale) and BMI in HC and UD.

### Behavioral

#### Number of pleasant responses and RT for food vs. object items

Participants gave more ‘pleasant’ responses food vs. objects (pleasant food: 78.6(2.16)%, pleasant objects: 58.5(2.16)%; F(1,88)=79.5, p<0.001) and had faster RT for *pleasant* food items compared to *pleasant* objects (food: 800(2)ms, objects: 890(2)ms), F(1,88)=103, p<0.001). These effects did not differ by participants’ diagnosis.

#### Rating of food items

The percent of food items that participants rated as pleasant during the scan significantly correlated with the mean ‘LIKED’ ratings (r=0.39, p=0.0001), but did not correlate with the mean ‘WANT’ ratings (r=0.04, p>0.1). The mean ‘LIKED’ or ‘WANTED’ ratings did not depend on the UD/HC status, BMI, lifetime depression severity, or BMI-by-depression interaction.

### Neuroimaging

A significant BMI-by-lifetime depression severity interaction effect on brain activation during *anticipation* of food vs. object pictures was observed only in the right inferior frontal gyrus (RIFG) pars opercularis (nvox=31, z-max=5.06, [60,14,10]; Figure 3). Figure 3A shows the inclusive mask (in tan color) that included the voxels that increased in activation during *anticipation* and/or pleasant/unpleasant *rating* of food and object items. The RIFG region is shown in purple on the central image. Figure 3B illustrates the BMI-by-lifetime depression severity interaction with the colored dots representing each data point. Darker blue color reflects greater activation during object vs. food *anticipation*, while darker red color reflects greater activation during food vs object *anticipation*. Figure 3C illustrates the same interaction but separately for HC and UD. The follow-up analyses that tested the BMI*lifetime depression severity interaction effect on the percent signal changes in the RIFG during food vs. object *anticipation* separately in UD and HC, showed a significant interaction effect in UD (t=-3.16, p= 0.003), but not in HC (t=0.94, p= 0.36).

**Figure 3.**
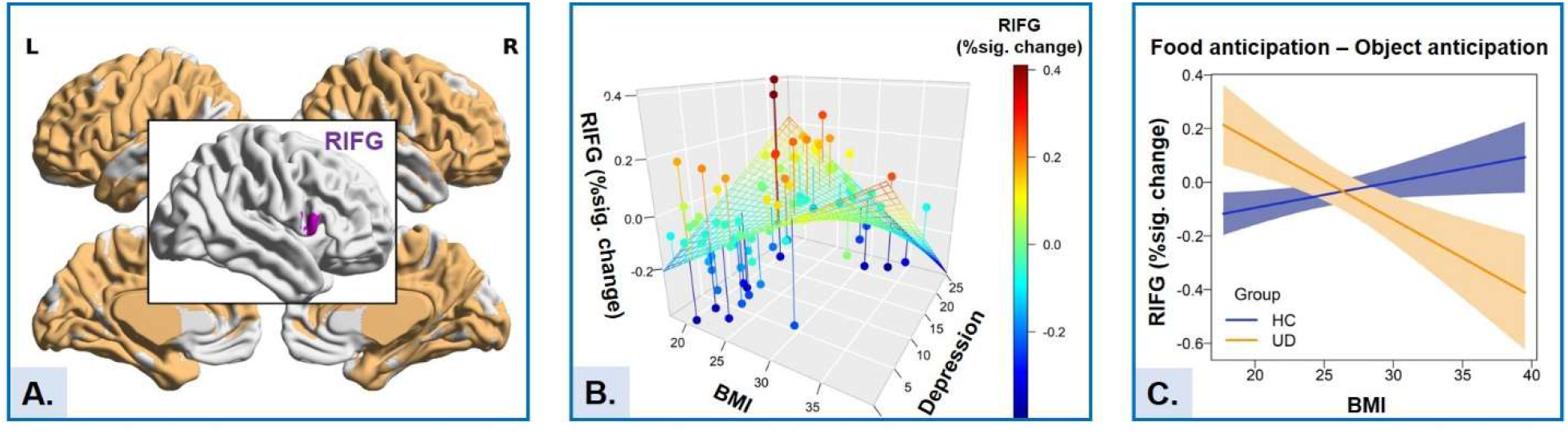
BMI by lifetime depression severity interaction effect on activation in the right inferior frontal gyrus (RIFG). (A) RIFG activation (in purple) and the inclusive activation map (in tan) for all anticipation and all processing trials. (B) BMI by lifetime depression severity interaction effect on the RIFG activation. (C) BMI by Group interaction effect on the RIFG activation

When we controlled age, sex, and IQ on the relationship between the difference in the RIFG activation during food vs. object *anticipation* and the difference in the RIFG activation during food vs. object item *pleasant/unpleasant ratings*, we found a significant negative partial correlation in UD (r=-0.71, p<0.001), HC (r=-0.57, p<0.001), and the whole sample (r=-0.63, p<0.001) (Figure 4A).

**Figure 4.**
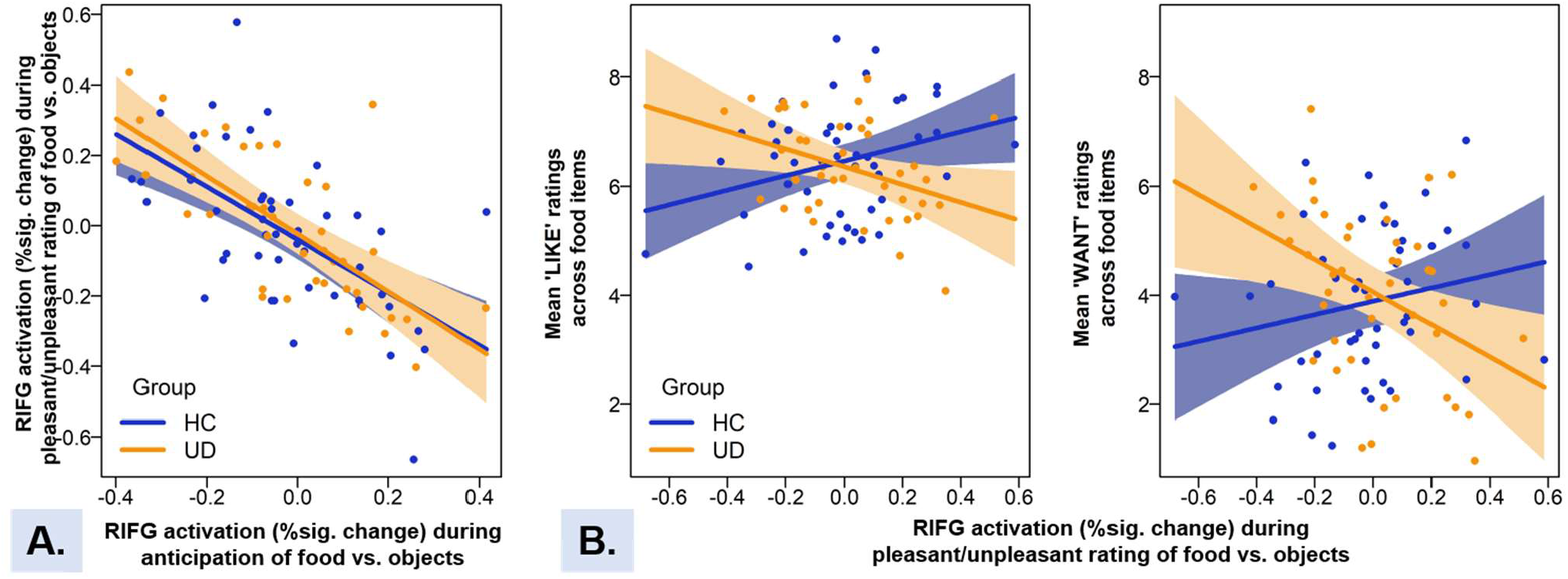
(A) Correlation between the differences in food vs. object *anticipation* and food vs. object *pleasant/unpleasant rating* in the right inferior frontal gyrus (RIFG) in healthy controls (HC) and individuals with unipolar depression (UD). (B) The relationship between food vs object differences during *pleasant/unpleasant rating* in the RIFG and ‘LIKE’ and ‘WANT’ ratings in HC and UD.

The moderation analyses with the Bonferroni corrected significance level p=0.0125 showed that the relationship between percent signal changes in the RIFG for *pleasant/unpleasant rating* of food vs. objects and ‘LIKE’ or ‘WANT’ ratings significantly depended on UD/HC diagnostic status (‘LIKE’: t=-3.1, p=0.003; ‘WANT’: t=-2.88, p=0.005; Figure 4B). Further exploration of the moderation effect for the ‘LIKE’ responses showed that the effect was driven by the negative relation between these variables in UD (‘LIKE’: t=-2.4, p=0.02), but not HC (‘LIKE’: t=1.6, p=0.11). The moderation effect for the ‘WANT’ responses was driven by the negative relationship in UD (‘WANT’: t=-2.4, p=0.02), but the positive relationship in HC (‘WANT’: t=2.26, p=0.03).

The ***exploratory analyses*** conducted in UD revealed no significant correlation between the differences in the RIFG activation during anticipation of food vs. objects and medload (p=0.44), the number of psychotropic medications (p=0.3), or the number of episodes of depression (p=0.44). There was a marginally significant positive relationship between the differences in anticipation of food vs. objects and age of illness onset (r=0.3, p=0.053) with greater differences observed in UD whose depression onset occurred later in life.

## 4 DISCUSSION

In this study, we investigated for the first time how the interplay between BMI and dimensional lifetime depression symptom severity was associated with brain activation during *anticipation* and *pleasant/unpleasant rating* of food, compared to objects. The results of the study allowed us to develop the empirical model depicting how the interplay between BMI and depression may affect appetitive behavior in individuals with depressive disorders (Figure 5).

**Figure 5.**
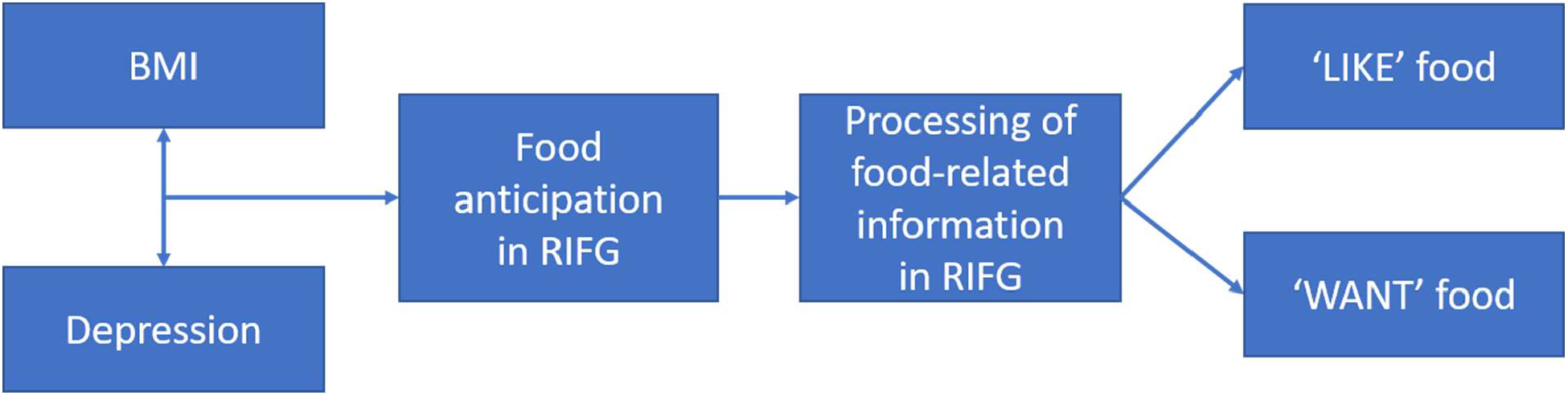
Empirical model of how BMI and depression interaction affects how much people like and want.

Consistent with the previous studies (3,5,6,8), we found a significant positive relationship between BMI and lifetime depression severity in UD: higher BMI was associated with more severe depression. No such association was found in HC. The interplay between BMI and lifetime depression was related to RIFG activation during food *anticipation* across all participants. This relationship was driven by the UD group in which higher BMI individuals with UD showed decreased RIFG activation (food<object) during *anticipation*, while normal BMI UD showed increased RIFG activation (food>objects) in the same condition. These results could not be explained by the differences in the processing of food stimuli by overweight/obese and normal weight individuals because there were no visual, olfactory, or other sensory food-related information during *anticipation*. Instead, the geometric shapes (a circle for objects and a triangle for food) were used to predict the upcoming stimulus category. The cognitive processing during this phase of the experiment was similar to a “I wish I could eat a steak right now” thought that one might have in the middle of a meeting despite the absence of food cues. These results also could not be explained by participants’ level of hunger/satiation (43) because HC and UD in our study did not differ in their hunger/satiation level prior to the scan.

Further exploration of the activation patterns in the RIFG showed that the magnitude of the food vs. object differences in the RIFG during *anticipation* was negatively correlated with the magnitude of the food vs. object differences in the RIFG during *pleasant/unpleasant rating*. Participants who showed more negative RIFG activation during *anticipation* (food<object) showed more positive RIFG activation during *pleasant/unpleasant rating* (food>object). The IFG is involved in emotion generation and regulation (44,45). The RIFG, specifically, is linked to response inhibition and attention control (46,47,49,51) that are necessary to control appetitive behavior. Our previous studies showed that the IFG significantly *increased* in activation during *performance* on a difficult, compared to an easy, working memory task, but significantly *decreased* in activation during *anticipation* of performing a difficult vs. easy working memory tasks (18,48). It was proposed that this effect may be related to controlling available cognitive resources by decreasing interference from the sources irrelevant to the current task (18,48). In the present study, a decrease in RIFG activation during food *anticipation* but an increase during *pleasant/unpleasant rating* of food observed in high BMI individuals with UD suggests that these individuals, compared to other study participants, face greater cognitive challenge during anticipating and processing of food-related information.

While preparing for and making pleasant/unpleasant judgments about food is not necessarily more difficult than making such judgments about objects, food judgments may be more emotional for overweight/obese individuals with UD. These individuals might also have difficulty to disengage from thinking about food when exposed to food stimuli perhaps out of concern that they may become over-invested in eating. High hedonic valence and emotional salience of food, compared to objects, in this study were supported by the findings that the judgments about food were made faster and were more pleasant than those about objects. These results are consistent with the findings of greater attentional bias to food stimuli in overweight/ obese individuals (50). Given that processing of emotional information requires more attentional resources (52,53), cognitive and neural mechanisms of mental preparation for processing of food stimuli may resemble the mechanisms of mental preparation for a difficult task. This is especially relevant to obese individuals with UD who decreased their RIFG activation during food anticipation the most.

Making pleasant/unpleasant judgments involves deep stimulus encoding (54,55). Based on this, we hypothesized that the responses and brain activation during *pleasant/unpleasant rating* of food items could be related to how much participants liked and wanted to eat those food items. We found that the diagnostic status moderated the relationship between RIFG activation for food vs. objects during *pleasant/unpleasant ratings*, but not during *anticipation*, and how much participants liked and wanted to eat food shown during the study. Surprisingly, participants with UD who showed higher RIFG activation for food during *pleasant/unpleasant rating* reported that they liked and wanted food less than participants with UD who showed lower RIFG activation. Given that the RIFG is involved in inhibitory control (46), these results may indicate that increased RIFG activation in UD may be related to an effort to inhibit processing of food images.

Previous studies have shown that the insula, posterior fusiform gyrus, and lateral orbitofrontal cortex activated in response to food vs. non-food items (56,57). The only recent study comparing obese individuals with and without major depressive disorder found stronger activation in the precuneus and anterior cingulate cortex in the former individuals during judging neutral words as pleasant or unpleasant (15). In addition, greater activation in insula, striatum and fusiform gyrus predicted less successful longitudinal outcome in the weight maintenance program (58). In this study, the RIFG was the only region that survived correction for multiple contrasts. However, results that were corrected for multiple comparisons at the brain level, but not for multiple contrasts, revealed several additional regions that included insula, orbital frontal, dorsal medial and lateral prefrontal, and anterior cingulate cortices (Supplemental Materials, Table 1S) that were reported in previous studies of obesity and food vs. non-food item processing. Even though our sample of 90 subjects is one of the largest in the relevant literature, the activation differences during task anticipation could be subtle and require even larger sample size.

The exploratory analyses showed the main outcomes of this study were unrelated to psychotropic medications load or the number of psychotropic medications in UD. These results are consistent with the recent findings that weight increase in mood disordered individuals is better explained by depression status rather than the use of antidepressants (59)

### Limitations

One limitation that may be important to consider in future work involves rating food stimuli as pleasant or unpleasant. Comparing brain activation for the food stimuli rated as pleasant vs. unpleasant would be potentially informative, but we were not able to examine this question: there were many more pleasant than unpleasant responses to food stimuli with many participants rated all food items as pleasant. A second limitation is that the VAS scores were only available for 60% of participants. A third limitation concerns the assessment of BMI. Body height and weight measurement equipment was not available in the laboratory, therefore we used self-reported height and weight. While overweight/obese people tend to underestimate the weight status of self and other people (60,61), the subjects in our study were not asked to estimate their weight status. They also were not aware that BMI might be of interest for the study except to determine MRI eligibility. Therefore, we hope that the body size was reported accurately. Finally, we were not able to recruit individuals with extreme obesity (BMI>40) due to the scanner weight limit and the size of the radio frequency coil. These limitations can be overcome by employing functional near infrared spectroscopy (fNIRS) that has recently been successfully implemented in the obesity (62,63) and depression (64) studies.

*In summary*, this study showed that the interplay between depression and obesity in individuals with UD affects activation patterns in the RIFG during food *anticipation*. Most symptomatic and overweight/obese individuals with UD showed decreased RIFG activation during food *anticipation*, while individuals with UD who were less symptomatic and overweight/obese, or symptomatic but had normal BMI showed increased RIFG activation during food *anticipation*. As a*nticipatory* RIFG activation was negatively associated with RIFG activation during *pleasant/unpleasant rating* of food items, overweight/obese individuals with UD increased RIFG activation during *pleasant/unpleasant rating* of food. One would expect that overweight/obese individuals would have higher ‘LIKE’ and ‘WANT’ ratings because of attentional bias toward food in these individuals (50). However, individuals with UD who increased RIFG activation during *pleasant/unpleasant* food rating had lower ‘LIKE’ and ‘WANT’ scores than participants who decreased RIFG activation in this condition suggesting that they try to inhibit or otherwise control their bias toward food thus decreasing subjective feeling of how much they like or want to eat it. Given the frequency with which people encounter food and food cues, inhibiting natural cognitive and emotional response to food may be computationally and cognitively expensive and stressful, thus causing worsening of depression. We propose that the RIFG response to food cues may be a risk factor for both weight gain and worsening of depression in individuals with UD. Taking these findings into consideration during the development of intervention strategies may improve effectiveness of interventions targeting both weight loss and depression improvement.

## FUNDING ACKNOWLEDGEMENTS

This work was supported by grants from the National Institute of Health R01MH114870 to A.M. and P41EB019936 to the Center for Reproducible Neuroimaging Computation (PI: Kennedy)

## Data Availability

Raw neuroimaging data will be available at OpenNeuro.
Other data will be deposited on the OSF and git repository.

## ACKNOWLEDGMENTS

The authors thank participants for taking part in this research study. We also thank Dr. Mary L. Phillips for fruitful discussions of the study design.

## AUTHORS CONTRIBUTION

A.M. – obtained funding, designed the study, acquired data, evaluated data quality, analyzed, and interpreted the data, drafted, and critically evaluated the manuscript

Y.O.H. – curated data organization and analyses, drafted and critically evaluated the manuscript S.S., R.R., M.L. – acquired data, evaluated data quality, drafted, and critically evaluated the manuscript

H.A.S. – curated participants’ recruitment, interpreted the data, critically evaluated the manuscript.

M.D.L. – curated study development, interpreted the data, critically evaluated the manuscript

All authors have read and approved the final version of the manuscript and agreed to be accountable for all aspects of this work.

